# The Impact of Technology-Enabled Virtual Medical Nutrition Therapy on Weight Loss in Adults with Overweight and Obesity: A Real-world, Observational Study

**DOI:** 10.1101/2024.12.17.24318819

**Authors:** Emily A. Hu, Tommy Kelley, Ajay Haryani

## Abstract

**Background:** Obesity represents a major public health crisis in the United States, imposing substantial health risks and economic costs. Medical nutrition therapy (MNT) has been demonstrated to be an effective intervention for weight loss among patients with obesity. With the rise of telehealth, virtual MNT has gained popularity as an accessible alternative to traditional in-person care. A nationwide program was developed that integrates virtual MNT with a companion mobile app, offering a comprehensive approach to weight management. However, the effectiveness of this combined intervention has not yet been evaluated.

**Objective:** To evaluate the effect of a virtual MNT program with a companion mobile app on weight loss in adults with overweight and obesity.

**Methods:** This retrospective cohort study included users of Nourish, a virtual MNT program with a companion mobile app, who attended at least one appointment between August 2023 and October 2024 and had a baseline body mass index (BMI) ≥30 kg/m² or a BMI between 27-30 kg/m² with diabetes or prediabetes. Engagement was assessed based on completed appointments and app usage. Mean weight change, mean percent weight change, and proportion of participants who achieved at least 3% and 5% weight loss were calculated. *T*-tests were used to evaluate the statistical significance of weight change. Subgroup analyses were performed by follow-up time between weights, number of appointments completed, and level of engagement by appointments and app usage.

**Results:** In total, 3,951 participants were included in the analysis. The mean (standard deviation (SD)) age was 38 (10) years, and 78% of participants were female. Weight loss was reported as a program goal by 70% of participants, with 31% self-reporting diabetes or prediabetes and 24% self-reporting a cardiovascular condition. Over a median follow-up of 2.2 months, 74% of participants experienced weight loss, and 34% and 17% achieved at least 3% and 5% weight loss, respectively. The mean (SD) weight change was -4.5 (8.9) pounds, corresponding to -2.0% (3.9) weight change (*P*<.001). Males and participants aged 60 years or older experienced greater weight loss than females and younger participants. Longer follow-up time between weights and a higher number of completed appointments (≥5 appointments) were significantly associated with a higher likelihood of achieving at least 5% weight loss (*P*<.001 for both). Additionally, participants who had the highest number of appointments and highest app engagement were more likely to achieve at least 5% weight loss compared to those with fewer appointments or lower app engagement (*P*<.001).

**Conclusions:** Enrollment in a virtual MNT program with a companion mobile app was associated with clinically meaningful weight loss among adults with overweight and obesity across the United States, highlighting the potential of tech-enabled solutions to provide scalable weight management strategies.

## Introduction

Obesity remains a critical public health challenge nationwide, affecting over 100 million adults across the United States (US) according to recent estimates (1). In the US, the prevalence of obesity has continued to rise, with over 40% of adults classified as obese in 2023 (2) and rates expected to exceed 60% in 2050, posing a significant public health threat given its impact on the existing chronic disease epidemic (3). This trend is associated with substantial health and economic burden, as obesity-related conditions, such as diabetes and other cardiovascular disease risk factors, are projected to worsen cardiovascular health and cost up to $1.49 trillion annually, along with additional losses from reduced productivity (4).

A cornerstone of obesity prevention and treatment is optimization of behavioral and lifestyle modifications, including adherence to regular physical activity and a well-balanced, heart-healthy diet (5). Effective nutritional counseling, as part of a medical nutrition program, has repeatedly proven to result in weight loss, thereby lowering an individual’s risk of obesity-related comorbidities, including diabetes and cardiovascular disease (6–10).

Medical nutrition therapy (MNT) is a treatment modality where a registered dietitian (RD) meets one-on-one with a patient to provide individualized lifestyle recommendations. MNT can be used to address a wide range of health conditions and may occur in various practice settings, such as outpatient and inpatient facilities. Following an initial intake, an RD meets with a patient regularly to individualize their treatment plan and help patients achieve their health goals. Data has consistently demonstrated the impact of MNT on reducing body weight, body mass index (BMI), and waist circumference and has been shown to improve weight-related risk factors, including hypertension, diabetes, and hyperlipidemia, and quality of life (6, 8). Despite its effectiveness, the utilization of MNT in practice is low relative to the prevalence of obesity for multiple reasons, including access, affordability, and awareness of this resource (11, 12).

During the COVID-19 pandemic, the adoption of telehealth significantly broadened healthcare accessibility (13). In response, the American Heart Association (AHA) issued a policy statement emphasizing the importance of expanding access to and optimizing the use of MNT, citing its numerous benefits for managing chronic diseases (14). Virtual MNT, which emerged as an opportunity to bridge the gap highlighted by the AHA, had the potential to not only improve access and convenience of MNT, but to also help achieve clinical goals. Emerging research has validated the potential of virtual MNT to deliver on the AHA mission by improving access, and in fact has resulted in better engagement and adherence resulting from the increased convenience of telehealth (15).

Starting in 2021, a national program, which combines virtual MNT with a companion mobile app, was launched to provide care for individuals with overweight and obesity seeking to improve their health. The app includes features such as meal logging, meal planning, and biometric tracking, which have been demonstrated to result in meaningful weight loss among patients with obesity in standalone mobile apps (16–18). By combining two proven interventions for weight loss, MNT and a companion mobile app, the program seeks to deepen patient engagement with their MNT treatment, and in turn see meaningful clinical outcomes.

To date, no studies have evaluated the effectiveness of this combined approach of virtual MNT with a companion mobile app. This study sought to evaluate the real-world impact of this combined solution on weight management among adults with overweight and obesity across the US.

## Methods

### Virtual MNT Program with a Companion Mobile App

Nourish is a national telehealth provider of virtual MNT delivered by a licensed RD, paired with a companion mobile app to offer asynchronous support and tools. Nourish has a network of over 1,500 RDs covering all major specialties, including overweight and obesity, diabetes, and heart disease, and can serve patients across all 50 states. Patients meet virtually in one-on-one sessions with their RD for an initial intake assessment. The RD then develops a personalized treatment plan focused on nutrition and lifestyle recommendations to help patients achieve their health goals. One-hour sessions are scheduled every one to three weeks, depending on the treatment plan. In addition to virtual appointments, patients also have access to a proprietary mobile app (Android, iOS) and web portal with features designed to complement the clinical intervention and increase engagement. Features include a self-reported health questionnaire, biometric progress tracking, meal logging, recipe and meal planning content, and on-demand chat with an RD (**Figure 1**).

**Figure 1.**
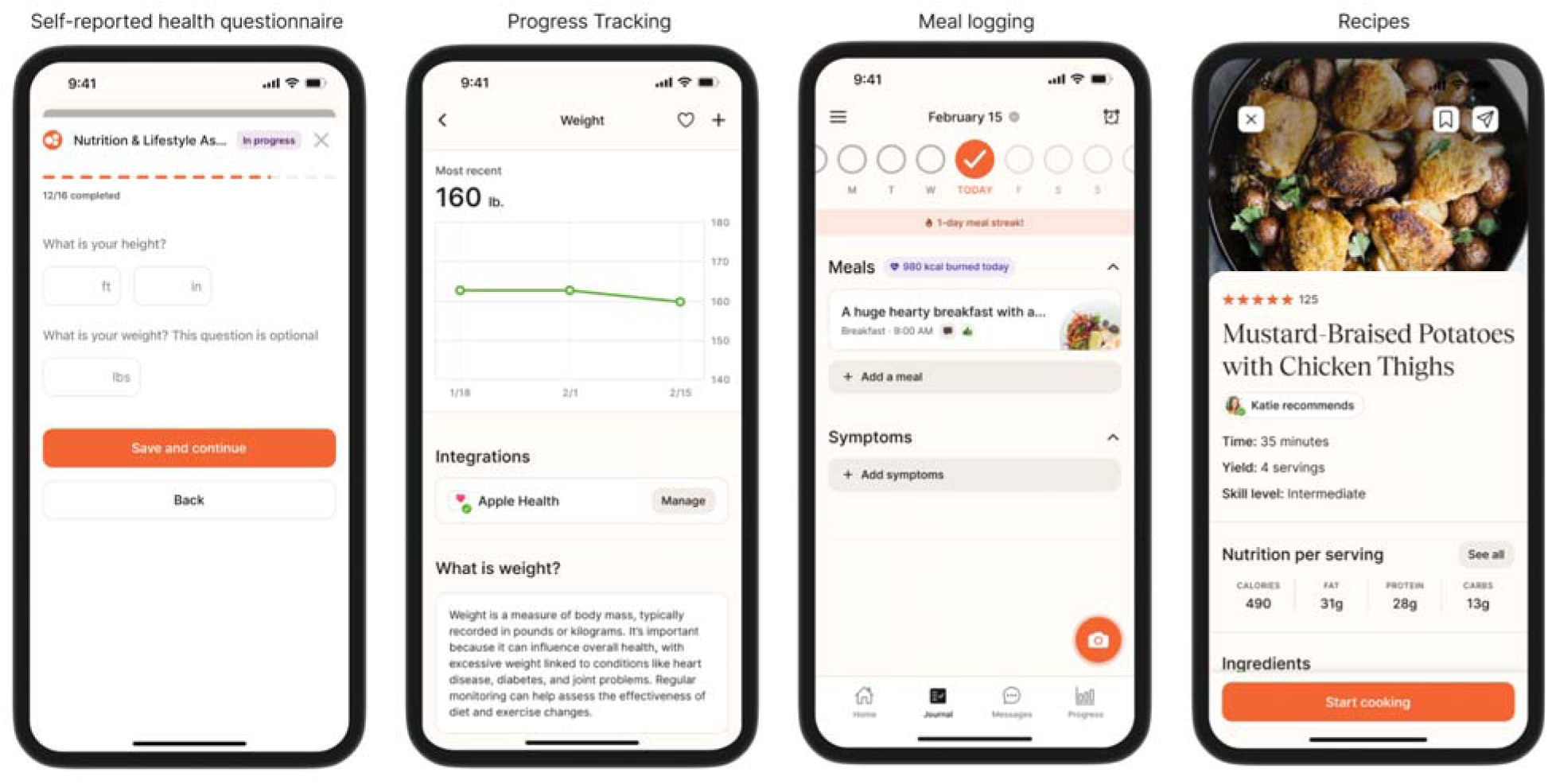
Screenshots of Nourish mobile app.

### Study Population

This retrospective cohort study evaluated weight change and engagement among users of a virtual MNT program with a companion mobile app who completed at least one appointment with an RD between August 2023 and October 2024. Participants were aged 18 years or older and met one of the following criteria: a BMI ≥30 kg/m^2^ or a BMI between 27 and 30 kg/m^2^ with self-reported diabetes or prediabetes. Exclusion criteria included reported usage of weight loss medications, history of bariatric surgery, or history of thyroid disorders. Participants were required to have at least two non-identical, self-reported weight measurements at least 30 days apart.

Baseline weight was defined as the earliest recorded weight within two weeks of the participant’s first appointment. The follow-up weight was the last recorded weight within 90 days of their last appointment, provided that at least four weeks had elapsed since the first appointment.

### Assessments

Participants were prompted to enter demographic and clinical information when they enrolled in the program. These self-reported assessments included sex, age, weight, height, goals for the program, and comorbidities such as diabetes or prediabetes and cardiovascular conditions such as high blood pressure and high cholesterol. Participants could update their weight at any time through the app, and RDs were prompted to record weight during appointments if participants did not update them in the app. All fields were optional except for date of birth and primary goal for enrolling in the program.

Based on self-reported height and weight data, BMI was calculated. Weight change was calculated as the difference between baseline weight and follow-up weight. Percent weight change was calculated as weight change divided by baseline weight. Follow-up time between weights was defined as days between the date that baseline weight was recorded and the date that follow-up weight was recorded. Additionally, follow-up time between appointments for participants who attended at least two appointments was calculated. The primary outcomes assessed were mean weight change (in pounds) and the proportion of participants who achieved at least 5% weight loss. Secondary weight outcomes included mean percent weight change and the proportion of participants who achieved any weight loss, at least 3%, at least 7%, and at least 10% weight loss.

To examine the relationship between engagement and weight reduction, engagement metrics based on app-generated data were also analyzed. Engagement was assessed based on the number of completed appointments and the number of total touchpoints with the app. The number of touchpoints with the app was calculated based on total engagements with key app features, including number of meals logged, number of messages sent to their RD, and number of biometric values (e.g., weight, height, blood pressure, cholesterol) entered.

### Statistical Analyses

Baseline characteristics were summarized for the total population and further stratified by whether participants achieved at least 5% weight loss. Descriptive statistics, including means, standard deviations (SD), medians, interquartile ranges (IQR), and ranges for continuous variables and percentages for categorical variables were reported. Group comparisons were conducted using analysis of variance (ANOVA) for continuous variables and chi-square tests for categorical variables. *T*-tests were conducted to determine whether weight changes significantly differed from zero.

The mean (SD) weight change and mean (SD) percent weight change were calculated for the total population and stratified by whether participants achieved at least 5% weight loss. Additionally, mean (SD) weight change, mean (SD) percent weight change, and the percent of participants who achieved at least 5% weight loss were stratified by baseline characteristics. ANOVA and chi-square tests were used to test for significant differences within each subgroup.

The proportion of participants achieving at least 3% and 5% weight loss were analyzed according to follow-up time between weights (<2 months, 2 to up to 3 months, 3 to up to 6 months, and ≥6 months) and the number of appointments completed (1-2, 3-4, 5-7, and ≥8 appointments). Further, the proportion of participants who achieved any and at least 3%, 5%, 7%, and 10% weight loss was calculated among the total population and by whether participants completed at least 5 appointments (the mean number of appointments in the study population). To evaluate the potential interaction of appointments and app touchpoints on weight loss, participants were stratified by combined appointment engagement and app usage. The mean values were used as the basis to define four subgroups of high or low appointment frequency (mean of 5 appointments) and app usage (mean of 100 touchpoints, approximating 9 touchpoints per week based on a mean follow-up time of 80 days). Highest engagement was defined as ≥5 appointments and ≥100 touchpoints with the app. Lowest engagement was defined as <5 appointments and <100 touchpoints with the app.

A *P* value less than .05 was considered to be statistically significant for all tests. Stata version 18 (StataCorp) was used for all analyses.

The study was declared exempt from institutional review board oversight by Advarra Institutional Review Board given the retrospective design of the study and less than minimal risk to participants.

## Results

### Participant characteristics

In total, 3,951 participants were included in this analysis. Baseline characteristics are presented in **Table 1**. Participants had a mean (SD) age of 38 (10) years, mean (SD) BMI of 37 (6) kg/m^2^, and were primarily female (78%). About half of participants had a BMI greater than 35 kg/m^2^ and a majority (70%) reported that weight loss was a goal for them. Diabetes or prediabetes and cardiovascular conditions were reported among 31% and 24% of participants, respectively. The mean (SD) and median (IQR) follow-up time between weight entries was 80 (49) days and 67 (47-95) days, respectively. The mean (SD) and median (IQR) time between appointments was 70 (61) days and 52 (26-95) days. In total, participants attended a total of 18,966 appointments (mean [SD]: 4.8 [4.0]) and had 393,131 touchpoints with the app (mean [SD]: 100 [101]) (data not shown).

**Table 1.** Baseline characteristics.

|  | **Total Population**<br>**(N=3,951)** | **Achieved ≥5%**<br>**weight loss**<br>**(n=689)** | **Achieved <5%**<br>**weight loss**<br>**(n=3,262)** | ***P*** |
| --- | --- | --- | --- | --- |
| Sex<sup>a</sup>, n (%) |  |  |  |  |
| Male | 759 (19%) | 151 (22%) | 608 (19%) | .05 |
| Female | 3,082 (78%) | 514 (75%) | 2,568 (79%) |  |
| Age<sup>a</sup>, years |  |  |  |  |
| Mean (SD) | 38 (10) | 38 (11) | 38 (10) | .02 |
| Median (IQR) | 36 (30-44) | 36 (31-44) | 36 (30-43) |  |
| Range | 18-78 | 19-78 | 18-78 |  |
| Age group, n (%) |  |  |  |  |
| 18-29 years | 809 (20%) | 138 (20%) | 671 (21%) | .03 |
| 30-39 years | 1,653 (42%) | 280 (41%) | 1,373 (42%) |  |
| 40-59 years | 1,237 (31%) | 209 (30%) | 1,028 (32%) |  |
| ≥60 years | 141 (4%) | 38 (6%) | 103 (3%) |  |
| Follow-up time<br>between weights, days |  |  |  |  |
| Mean (SD) | 80 (49) | 97 (53) | 76 (47) | <.001 |
| Median (IQR) | 67 (47-95) | 85 (62-116) | 63 (45-90) |  |
| Range | 30-424 | 30-403 | 30-424 |  |
| Follow-up time<br>between<br>appointments<sup>b</sup>, days |  |  |  |  |
| Mean (SD) | 70 (61) | 81 (67) | 67 (60) | .002 |
| Median (IQR) | 52 (26-95) | 64 (29-112) | 49 (24-91) |  |
| Range | 1-412 | 3-361 | 1-412 |  |
| Weight, pounds |  |  |  |  |
| Mean (SD) | 228 (44) | 229 (44) | 227 (44) | .4 |
| Median (IQR) | 220 (195-252) | 220 (196-255) | 220 (195-252) |  |
| Range | 131-398 | 142-396 | 131-398 |  |
| BMI, kg/m<sup>2</sup> |  |  |  |  |
| Mean (SD) | 37 (6) | 37 (6) | 37 (6) | .4 |
| Median (IQR) | 35 (32-40) | 36 (32-40) | 35 (32-40) |  |
| Range | 27-75 | 27-75 | 27-67 |  |
| Baseline BMI, n (%) |  |  |  |  |
| 27-<30 kg/m<sup>2</sup> | 181 (5%) | 28 (4%) | 153 (5%) | .8 |
| 30-<35 kg/m<sup>2</sup> | 1,704 (43%) | 287 (42%) | 1,417 (43%) |  |
| 35-<40 kg/m<sup>2</sup> | 1,048 (27%) | 193 (28%) | 855 (26%) |  |
| 40-<50 kg/m<sup>2</sup> | 834 (21%) | 149 (22%) | 685 (21%) |  |
| ≥50 kg/m<sup>2</sup> | 184 (5%) | 32 (5%) | 152 (5%) |  |
| Diabetes or<br>prediabetes, n (%) | 1,204 (31%) | 227 (33%) | 977 (30%) | .1 |
| Cardiovascular<br>condition, n (%) | 939 (24%) | 168 (24%) | 771 (24%) | .7 |
| Weight loss goal, n (%) | 2,748 (70%) | 487 (71%) | 2,261 (69%) | .5 |

Abbreviations: BMI, body mass index; IQR, interquartile range; kg/m<sup>2</sup>, kilograms per meter squared; SD, standard deviation
<sup>a</sup> 3% (n=151) of the total population did not report sex nor age and were not included in summaries of sex and age.
<sup>b</sup> Among participants who had at least two appointments (n=3,269).

A greater proportion of participants who achieved at least 5% weight loss were male (*P*=0.05) and aged 60 years or older (*P*=0.03). Although not statistically significant, participants who self-reported diabetes or prediabetes were more likely to achieve at least 5% weight loss.

### Weight change

The mean (SD) weight change was -4.5 (8.9) pounds (*P*<.001) and the mean (SD) percent weight change was -2.0% (3.9) (*P*<.001) (**Table 2**). Among those who achieved at least 5% weight loss, the mean (SD) weight change was -17.6 (6.3) pounds (*P*<.001) and the mean (SD) percent weight change was -7.7% (2.4) (*P*<.001), while those who did not achieve at least 5% weight loss had a mean (SD) weight change of -1.8 (6.6) pounds and a mean (SD) percent weight change of -0.7% (3.0) (*P*<.001).

**Table 2.**
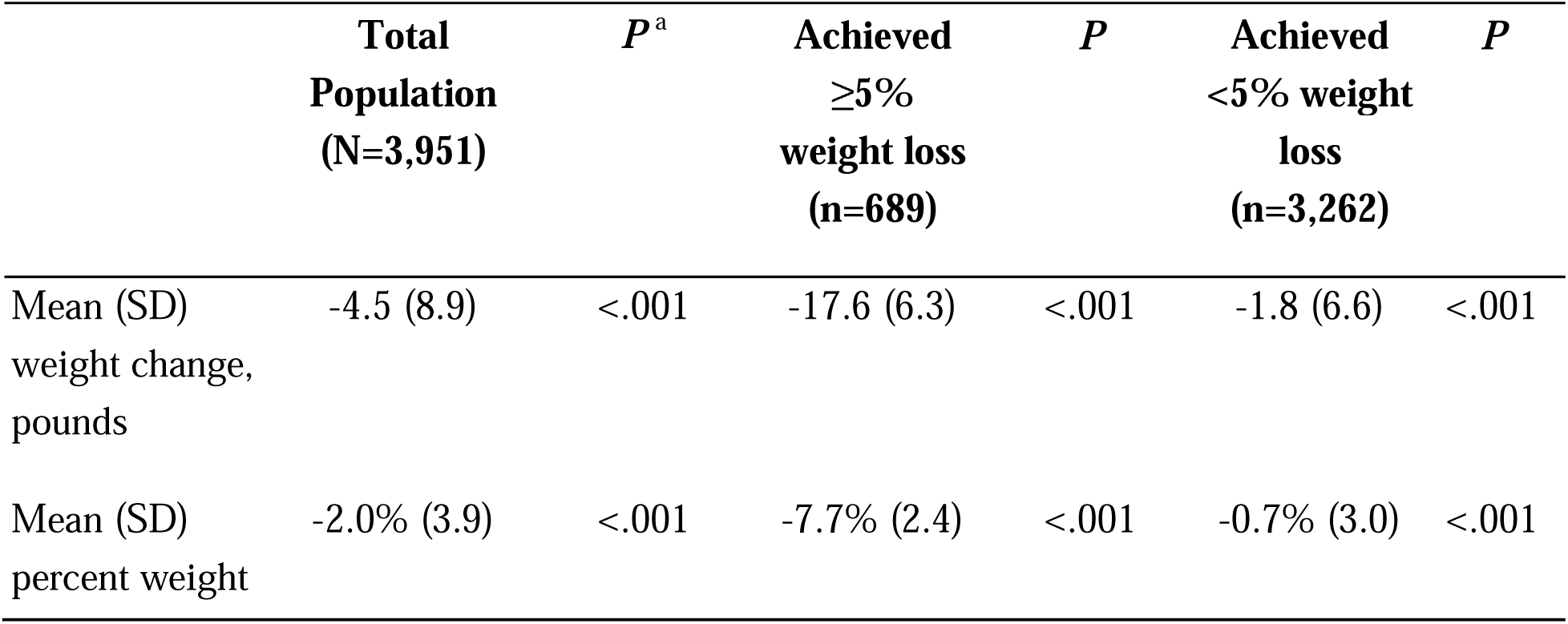
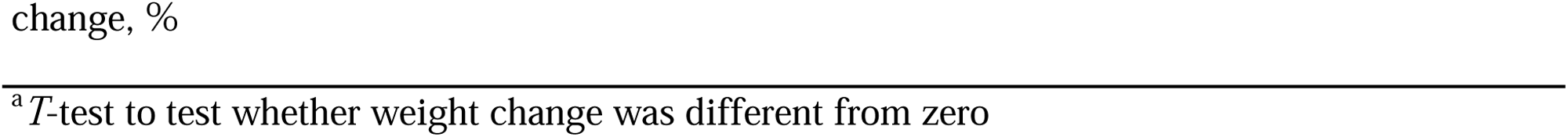
Mean weight change and percent weight change among the total population and stratified by 5% weight loss.

In total, 74% of participants lost any weight, 34% of participants achieved at least 3% weight loss, 17% achieved at least 5% weight loss, 9% achieved at least 7% weight loss, and 3% achieved at least 10% weight loss (**Supplemental Table**).

Table 3 summarizes mean (SD) weight change, mean (SD) percentage weight change, and the proportion of participants who achieved at least 5% weight loss, stratified by baseline characteristics. When comparing results by sex, males experienced a significantly greater mean (SD) weight change (-6.1 [9.4] pounds) compared to females (-4.1 [8.7] pounds) (*P*<.001). Baseline BMI also showed a strong association with weight loss, as participants with a BMI of 50 kg/m² or higher experienced significantly greater weight loss than those with a lower BMI (*P*<.001). Notably, participants who entered the program with a stated goal of losing weight experienced significantly more weight loss than those without this goal (*P*<.001). However, they were not more likely to achieve at least 5% weight loss.

**Table 3.**
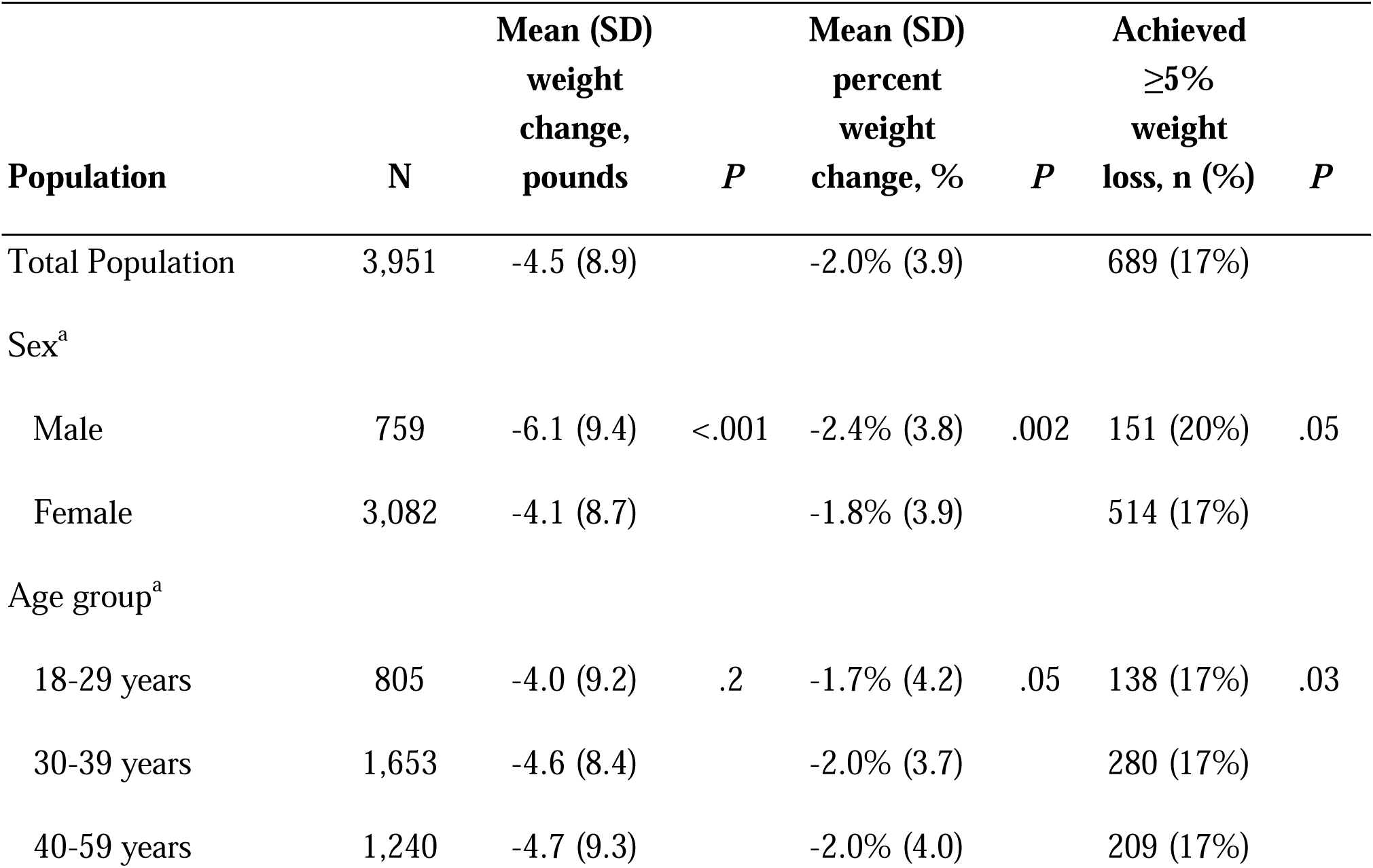
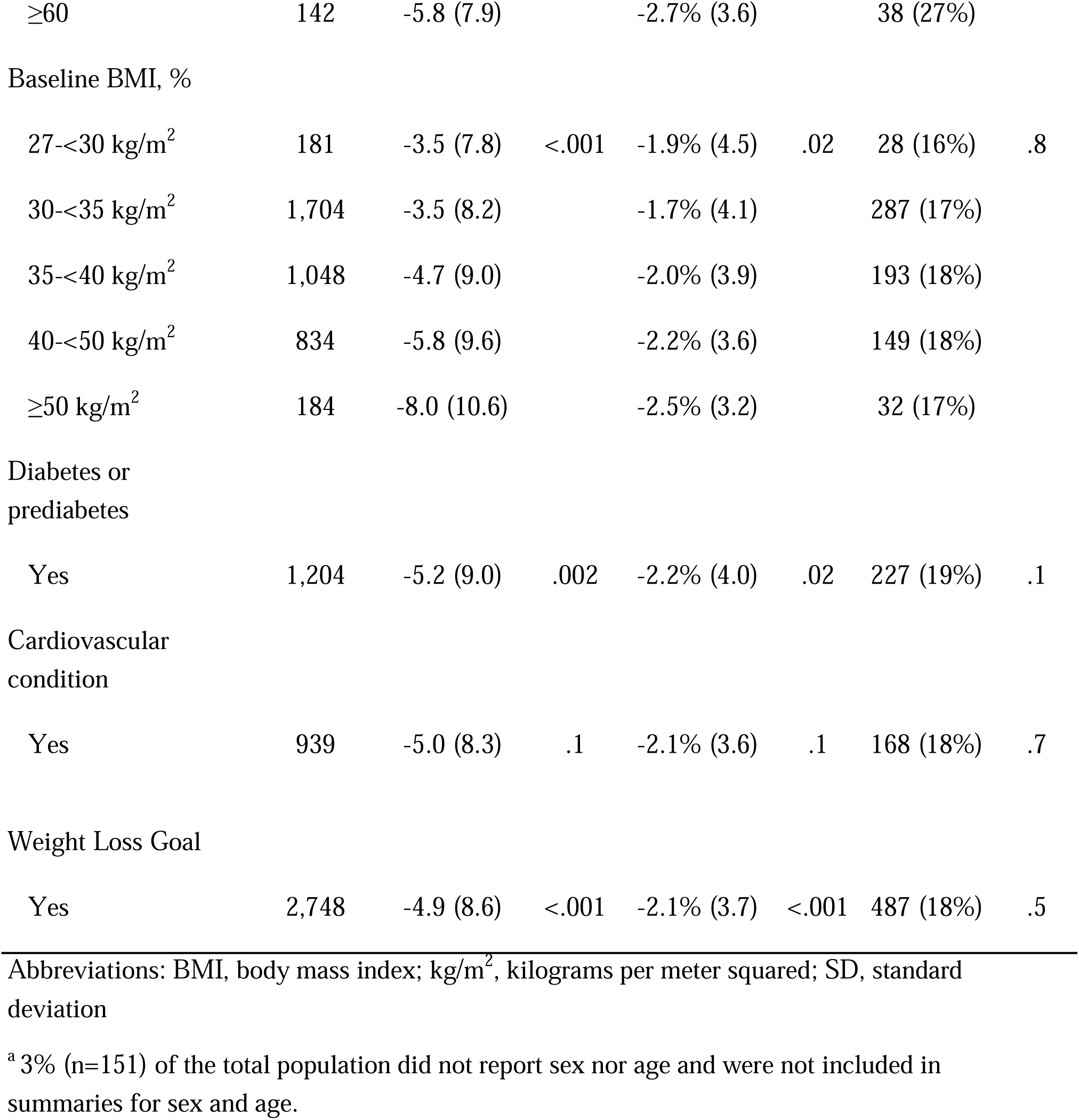
Mean weight change, percent weight change, and proportion of participants who achieved at least 5% weight loss by baseline characteristics.

### Engagement with Program

The proportion of participants who achieved at least 3% and 5% weight loss, stratified by time between weights, is presented in **Figure 2**. Participants who engaged with the program for longer periods of time were more likely to achieve at least 3% (*P*<.001) and 5% weight loss (*P*<.001).

**Figure 2.**
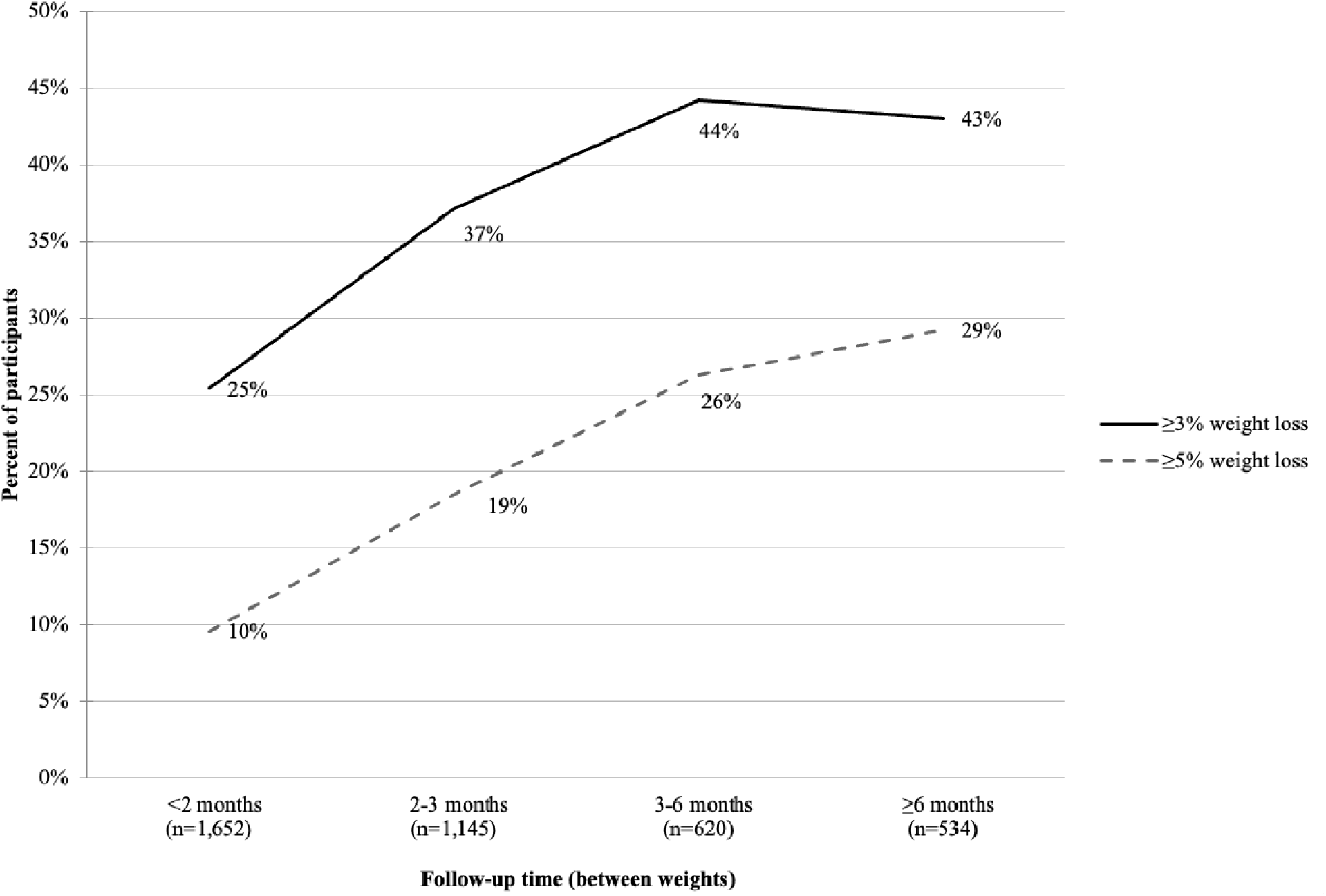
Percent of participants who achieved at least 3% and 5% weight loss according to follow-up time.

Figure 3 illustrates the proportion of participants achieving at least 3% and 5% weight loss, stratified by the number of completed appointments (1–2, 3–4, 5–7, and ≥8 appointments). Participants who attended more appointments had a significantly greater likelihood of achieving at least 3% weight loss (*P*<.001) and at least 5% weight loss (*P*<.001).

**Figure 3.**
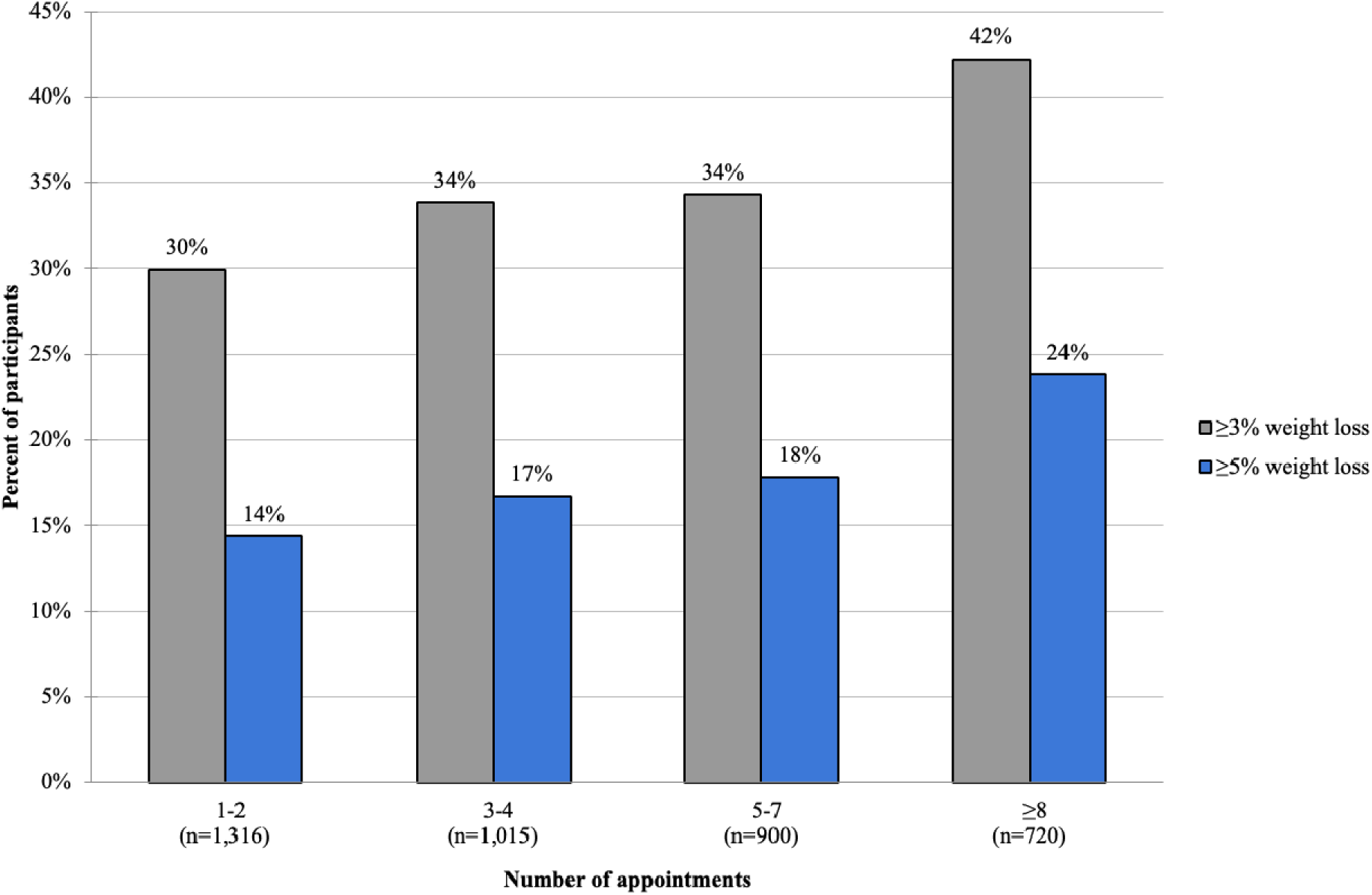
Percent of participants who achieved at least 3% and 5% weight loss by number of appointments.

Those who completed at least five appointments had significantly higher rates of achieving weight loss milestones compared to those who attended fewer than five appointments: 38% vs. 32% for at least 3% weight loss (*P*<.001), 20% vs. 15% for at least 5% weight loss (*P*<.001), 11% vs. 7% for at least 7% weight loss (*P*<.001), and 5% vs. 2% for at least 10% weight loss (*P*<.001) (**Supplemental Table**).

### Engagement with MNT and App

A significant interaction was observed between appointment completion and app engagement in predicting the likelihood of achieving at least 5% weight loss (*P*<.001; Figure 4). Participants with high engagement in both appointments and app usage had the greatest likelihood of achieving at least 5% weight loss. The likelihood of this occurrence followed in descending order by participants with a high number of appointments but low app usage, a low number of appointments but high app usage, and low appointment attendance and low app usage.

**Figure 4.**
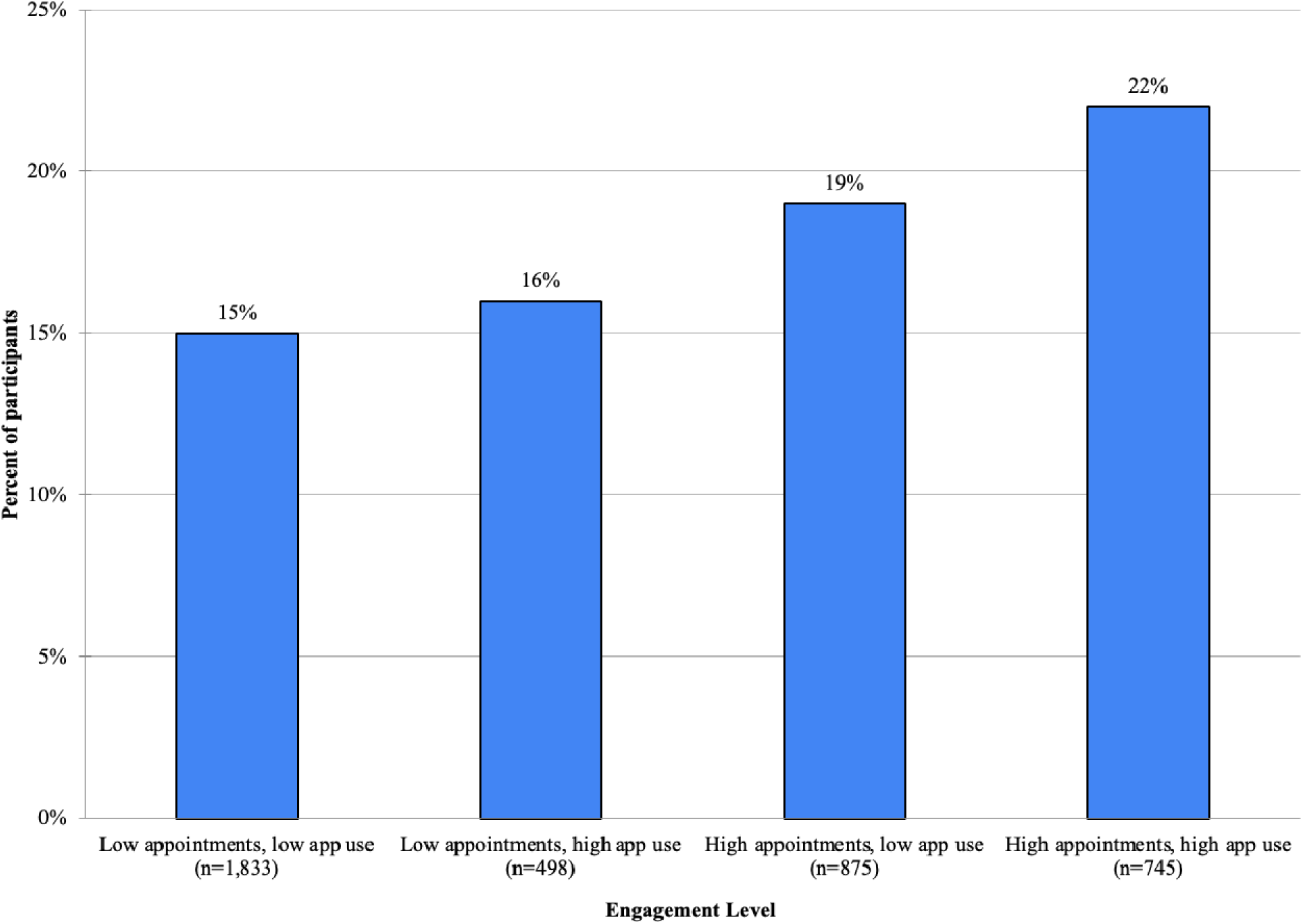
Percent of participants who achieved at least 5% weight loss by engagement data^a, b^. ^a^ Low appointments defined as <5 appointments; high appointments defined as ≥5 appointments using mean number of appointments; low app usage defined as used <100 touchpoints (9 touchpoints per week); high app usage defined as ≥100 touchpoints using the mean number of touchpoints ^b^ App usage includes meal logging, messages to RD, or health questionnaire entries

## Discussion

### Principal Findings

Among adults with overweight and obesity across the US, enrollment in a virtual MNT program with a companion mobile app was associated with clinically meaningful weight loss. Over a median of 2.2 months, the majority (74%) of participants lost weight and 17% of participants lost at least 5% of initial weight. The greatest weight loss was achieved by participants who were most engaged with both virtual MNT appointments and the mobile app, suggesting a significant benefit of integrating both resources into a nutritional therapy solution.

MNT has been proven to be an effective strategy to treat obesity by achieving significant weight randomized controlled trials comparing individualized nutrition care for weight management to usual care found a pooled mean change of -2.3 pounds over a median of 6 months (8). In the present study, participants who engaged with a virtual MNT program experienced a mean weight change of -4.5 pounds over a median of 2.2 months. Of note, several of the studies in the systematic review were focused on primary outcomes of blood pressure or diabetes, rather than weight loss. This suggests that when MNT is implemented with the primary goal of achieving weight loss, potentially more meaningful weight loss can be achieved.

Currently, MNT is covered broadly by private insurances for individuals with obesity, whereas Medicare and Medicaid coverage is more limited (19). There is also wide variability in the number of hours covered by payers for MNT, which is in part due to the fact that there is limited data regarding how to best optimize MNT with regards to appointment frequency and the impact of ongoing nutritional therapy for weight loss.

Based on the current analysis, while as few as one appointment resulted in meaningful weight loss, a higher number of appointments and longer time engaged in the program were directly correlated with a higher likelihood of clinically significant weight loss. In fact, the greatest outcomes were observed for participants who completed at least eight one-hour appointments. Therefore, in addition to prioritizing clinically effective MNT, an effective program must also emphasize long-term engagement and adherence to optimize clinically meaningful weight loss for individuals. In fact, this was substantiated by the Academy of Nutrition and Dietetics who reported that the most beneficial outcomes for blood pressure and fasting glucose resulted after at least 5 sessions with an RD over a period at least 12 months (20).

Although MNT helps individuals successfully lose weight, uptake has been low over the years, in part due to accessibility, affordability, and convenience (11, 12). At the same time, there has been a rise in digital health interventions for weight loss through mobile and web-based applications (21, 22). Several analyses evaluating the effectiveness of these commercial apps have been published and report meaningful weight loss among users in a real-world setting.

A retrospective cohort study of 35,921 participants with overweight and obesity who used a mobile app for weight loss, which included daily meal logging, activity monitoring, and personalized feedback, found that 78% of participants lost weight over a median of 8.8 months (17). While the present study had a smaller sample size, a comparable percentage of participants lost weight over a shorter time frame. One additional benefit of the current study compared to the study by Chin. et al is the inclusion of self-reported comorbidities and program goals, which allows for further understanding of how a weight loss program impacts patients by varying health statuses and program goals.

Similarly, among 8,977 adults with obesity, a digital nutrition platform with personalized recipes and online grocery tools resulted in a mean weight loss of 3.7 (1.5%) pounds over 11 months (16). In one-fifth the follow-up time, participants engaged with the current virtual MNT program with a companion mobile app achieved greater mean weight loss and mean percent weight loss. Furthermore, the current study includes detailed engagement data, whereas the previous study did not report engagement metrics; as a result, it remains unclear whether weight loss in the previous study was associated with active app use or passive engagement with the platform.

Recognizing the importance of a multi-faceted approach to weight loss and patients’ desire to have a mobile app or web-based platform to engage with during their weight loss journey, the present study aimed to better understand how app engagement further impacts weight loss success associated with MNT. The features of the mobile app component of this virtual MNT program were designed to drive engagement and facilitate consistent behavior change to ultimately result in better adherence to diet recommendations and greater weight loss. This has been validated in the analysis by Chin et al., where meal logging (odds ratio, OR: 10.69, 95% CI: 6.20-19.53; *P*<.001) and weight logging (OR: 0.59, 95% CI: 0.39-0.89; *P*<.001) were both associated with greater likelihood of successful weight loss (17).

Alternatively, the SMARTER trial evaluated the impact of a single 90-minute visit with an RD combined with an app for weight loss among individuals with overweight and obesity. The study concluded that the RD visit, along with app features such as meal logging and physical activity tracking via a Fitbit device, led to meaningful weight loss. However, they reported no additional benefit of receiving personalized, real-time feedback through in-app messages (23), which is in contrast to another study highlighting the benefit of an RD chat feature (24). Therefore, while the utility of each individual app-based feature requires additional investigation, multiple studies, including the current analysis, highlight that patients who additionally engaged with an app achieved the greatest weight loss.

### Limitations

There are several limitations of the current study to consider. First, this study is observational and does not include a control group. However, the real-world nature of this study provides valuable insights into the practical implementation and applicability of this solution across a broad patient population nationwide.

Second, self-reported data, including weights, are subject to recall bias, although previous research has suggested a moderate to strong correlation between self-reported weight data collected online and actual values (25). Future studies should aim to leverage objective clinical data to reduce bias.

Third, the follow-up time was short compared to other retrospective cohort studies. Despite this, similar weight loss was still achieved during the study period. Longer-term data are needed to assess the full effect and sustainability of weight loss achieved through this comprehensive solution.

Last, although the program covered all 50 states and likely represented a diverse patient population, the lack of comprehensive demographic data capture beyond age and sex limits the generalizability of this intervention and prevents assessment on the impact among individuals who are disproportionately affected by social determinants of health. Future research should explore the role of sociodemographic and cultural factors, including race, ethnicity, and geographic location (e.g., urban vs. rural settings), in health outcomes among users of this platform.

### Conclusions

The obesity epidemic in the US highlights the need for effective and comprehensive approaches to weight management. This study provides real-world evidence that engagement with a virtual MNT program and a companion mobile app is associated with clinically meaningful weight loss among individuals with overweight and obesity. It also highlights that combining virtual MNT with a companion mobile app may amplify the program’s effect on weight loss. These findings highlight the potential for virtual MNT programs with mobile apps to bridge the gap in access to effective nutrition care, offering a scalable and effective solution for weight management in individuals with overweight and obesity across the US.

**Supplemental Table.**
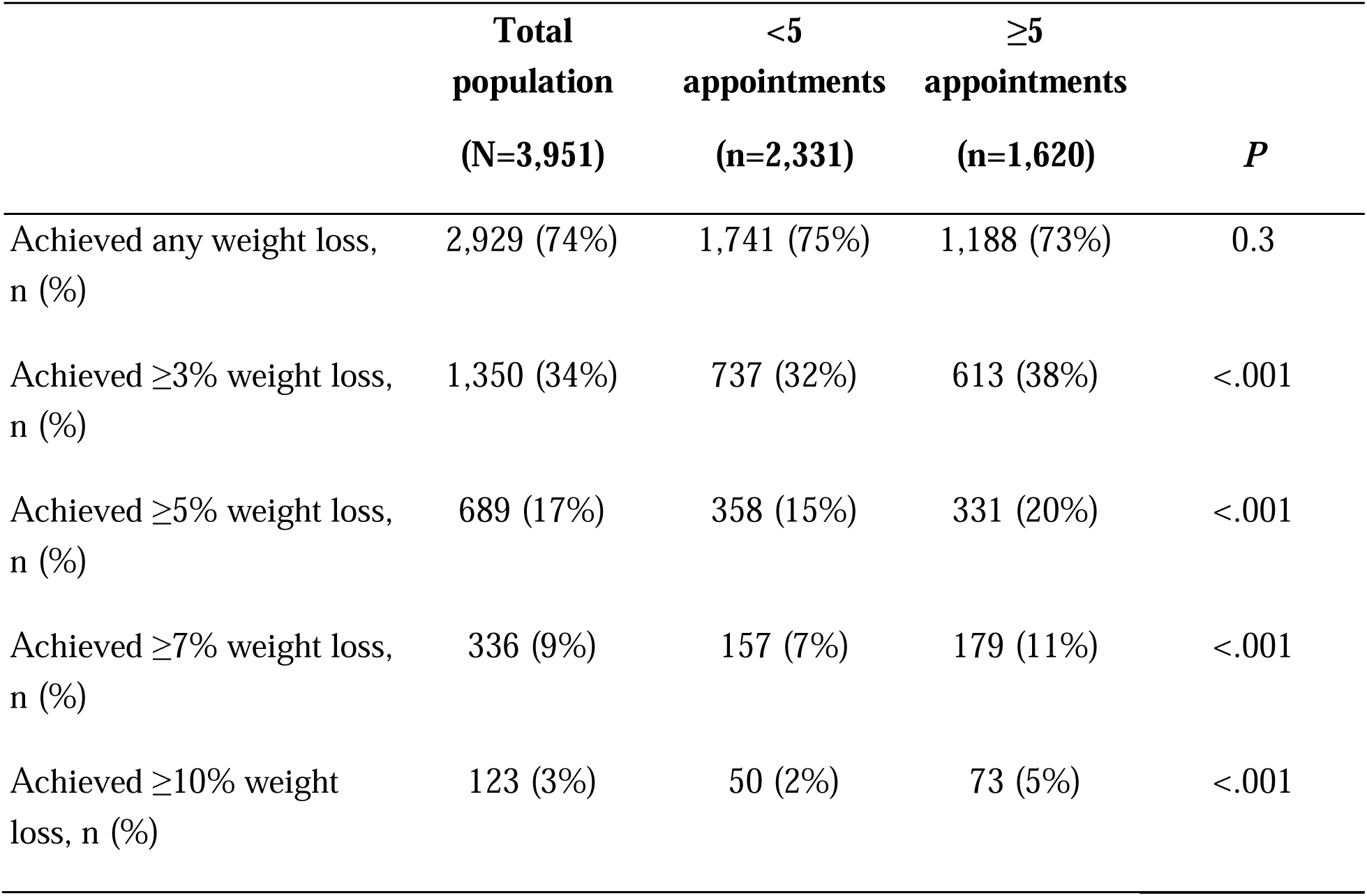
Percent of patients who achieved any and at least 3%, 5%, 7%, and 10% weight loss.

## Acknowledgements

The authors would like to acknowledge David Feldman and Julia Rayis for providing feedback and assistance with the manuscript.

## Data Availability

The data analyzed in this study are proprietary and cannot be publicly shared.

## Abbreviations

AHA: American Heart Association
ANOVA: analysis of variance
BMI: body mass index
IQR: interquartile range
OR: odds ratio
RD: registered dietitian
SD: standard deviation
US: United States

